# Disease burden outpaces essential diagnostic test availability for Neglected Tropical Diseases in India

**DOI:** 10.64898/2026.02.19.26346609

**Authors:** Zeeshan Mustafa, Biswajit Chakraborty, Amaj Ahmed Laskar, Rinki Kumar, Vipin Kumar, Kajal Arora, Anjam Hussain Barbhuiya, Leichombam Mohindro Singh, Kanu Shil, Sumit Roy, Mohammad Azam Khan, Mustafa A. Barbhuiya

**Author notes:** **Corresponding author:** Mustafa A. Barbhuiya, PhD Founder and Director, Foundation for Advancement of Essential Diagnostics Department of Pathology, Healthcare Delivery and Population Sciences UMass Chan Medical School- Baystate Regional Campus, Springfield, MA 01199, USA.

## Abstract

India carries a high burden of neglected tropical diseases (NTDs), yet the extent to which essential diagnostic services align with local disease burden across public health facilities remains unclear. We conducted a cross-sectional assessment of diagnostic availability for major NTDs in 332 public health facilities across seven states and one union territory, including sub-centers, primary health centers, community health centers, and district hospitals. Diagnostic availability for malaria, dengue, Japanese encephalitis, chikungunya, lymphatic Filariasis, Leishmaniasis, helminthic infections, and HIV was evaluated using the ICMR National Essential Diagnostics List (2019), and a Diagnostic Readiness Index (DRI) was calculated at the facility, district, and state levels. Big three diseases like tuberculosis, malaria, and HIV are not neglected anymore in terms of funding and research, but their interaction with classic NTDs highlights how these diseases, though different in classification, create a dual burden in vulnerable populations, like in India. Diagnostic readiness increased with higher levels of care but showed limited concordance with disease burden. Malaria diagnostics were widely available across all tiers (mean district DRI: 84.34%), reflecting sustained programmatic prioritization. In contrast, diagnostic availability for dengue (40.36%), lymphatic Filariasis (29.22%), helminthic infections (25.30%), Japanese encephalitis (8.13%), and Leishmaniasis (5.72%) remained low, including in districts reporting substantial disease burden. The greatest mismatch between burden and diagnostic availability was observed at sub-centers and primary health centers, whereas district hospitals showed a more favorable alignment. These findings indicate that essential diagnostic deployment for NTDs in India remains uneven and weakly responsive to epidemiological need. Strengthening burden-informed, decentralized access to point-of-care diagnostics—particularly at peripheral and primary care levels—is critical to improve early case detection, surveillance accuracy, and progress toward national NTD control and elimination targets.

**Author Summary:** Neglected tropical diseases (NTDs) continue to affect millions of people in India, particularly those living in underserved and resource-limited settings. Early and accurate diagnosis is essential for timely treatment, surveillance, and control; however, the availability of essential diagnostic services across different levels of the public health system remains poorly understood. In this study, we assessed the availability of diagnostics for major NTDs across 332 public health facilities, including sub-centers, primary health centers, community health centers, and district hospitals in seven states and one union territory, using the ICMR National Essential Diagnostics List (2019) and a Diagnostic Readiness Index.

We found that diagnostic readiness generally increased with higher levels of care but showed limited alignment with local disease burden. Malaria diagnostics were widely available, reflecting sustained programmatic prioritization, whereas diagnostics for dengue, lymphatic filariasis, helminthic infections, Japanese encephalitis, and leishmaniasis were markedly limited, even in areas with substantial reported burden. The largest gaps were observed at peripheral and primary care facilities, which are often the first point of contact for vulnerable populations.

Our findings highlight a critical mismatch between disease burden and diagnostic deployment for NTDs in India. Strengthening decentralized, burden-informed access to point-of-care diagnostics—especially at sub-centers and primary health centers—could improve early case detection, enhance surveillance accuracy, and support more effective control and elimination efforts. This study provides policy-relevant evidence to guide health system strengthening and equitable diagnostic access for NTD-endemic regions.

## Introduction

Neglected tropical diseases (NTDs) constitute a diverse group of infectious conditions that disproportionately affect populations living in poverty, particularly in tropical and subtropical regions. Characterized by their association with inadequate sanitation, limited access to healthcare, and fragile health systems, these diseases impose a substantial human and economic burden on affected communities (1). India, with its vast geographic, demographic, and socioeconomic diversity, is among the most affected countries globally. The World Health Organization (WHO) recognizes India as endemic for multiple high-burden NTDs, including vector-borne viral infections such as Dengue, Chikungunya, and Japanese encephalitis; parasitic diseases such as Malaria, Filariasis, Leishmaniasis, and Helminthic infections; and chronic infections such as HIV, which though not classified strictly as an NTD, intersects in epidemiology, morbidity, and health system challenges with NTDs in many low-resource settings (2–5).

The collective burden of these diseases is staggering. Dengue alone is estimated to affect over 33 million individuals annually in India, with cyclical outbreaks causing significant morbidity and mortality (6, 7). Chikungunya, once sporadic, has resurged since the early 2000s, leading to large-scale outbreaks associated with long-term joint disability and economic loss (8). Japanese encephalitis (JE), though less frequent, has one of the highest case-fatality rates among viral encephalitis, particularly in children, and remains endemic in parts of Andhra Pradesh, Assam, Bihar, Haryana, Karnataka, Kerala, Maharashtra, Manipur, Tamil Nadu, Odisha, Uttar Pradesh, and West Bengal (9–11). Malaria, although showing a decline under the National Vector Borne Disease Control Programme (NVBDCP), still accounts for more than 0.25 million reported cases annually (10). Lymphatic Filariasis (LF) continues to be a public health problem in 339 districts across 20 states/UTs, with more than 650 million Indians at risk of infection, many of whom already live with chronic disability and social stigma due to elephantiasis and hydrocele (10,12). Visceral leishmaniasis (kala-azar), concentrated in Bihar, Jharkhand, Uttar Pradesh, and West Bengal, remains one of the fatal parasitic diseases if left untreated, with India contributing nearly 18% of the global VL burden (13). Soil-transmitted helminthic (STH) infections, particularly roundworm, whipworm, and hookworm, disproportionately affect school-age children, impairing nutrition, growth, and cognitive development (14). Finally, HIV/AIDS, though subject to long-standing national control programs, no longer classified as neglected, continues to affect 2.4 million people in India, with co-infection risks and diagnostic challenges overlapping with NTDs (15). Despite their diverse etiologies, transmission dynamics, and clinical manifestations, these diseases share commonalities: they thrive in marginalized populations with limited healthcare access, contribute to cycles of poverty, and remain under-prioritized in comparison to other global health threats (16). One of the most critical barriers to effective prevention, control, and elimination of NTDs in India is the essential diagnostic availability gap. Early and accurate diagnosis is the foundation of disease control—enabling appropriate case management, outbreak detection, surveillance, and resource allocation. Yet, diagnostic services for NTDs in India remain patchy, with significant disparities across states, districts, and tiers of the health system (17–19). Despite inclusion in India’s NEDL, diagnostics for dengue, chikungunya, JE, Filariasis, kala-azar, and helminths remain scarce beyond tertiary centers. Malaria testing is wider but uneven in quality, while HIV diagnostics are stronger yet still limited in rural and remote settings. Diagnostic gaps cause delayed or missed diagnoses, increasing morbidity, mortality, and disability. They hinder outbreak detection, weaken surveillance, and misguide resource allocation. Limited diagnostic capacity also impedes the elimination of NTDs under the WHO’s 2021–2030 roadmap. At the community level, unavailability erodes trust, prompting patients to seek private or informal care, often with associated financial and treatment risks (20–22).

Addressing diagnostic challenges for neglected tropical diseases (NTDs) in India requires systematic evaluation. Although the National Essential Diagnostics List (NEDL) aims to standardize access, implementation remains uneven. This study assesses diagnostic availability for major NTDs—dengue, chikungunya, Japanese encephalitis, malaria, HIV, Filariasis, Leishmaniasis, and helminthic infections—across healthcare levels. By mapping diagnostic readiness against population burden, it identifies inequities, highlights bottlenecks, and proposes strategies to strengthen diagnostic systems for effective NTD control and universal health coverage.

## Methods

### Study design and setting

This cross-sectional descriptive study surveyed 332 health facilities between November 2023 and April 2025 across 24 districts in seven states and one union territory of India: Assam, Haryana, Himachal Pradesh, Manipur, Rajasthan, Tripura, Uttar Pradesh, and Delhi. The facilities surveyed included sub-centers (SCs), Primary Health Centers (PHCs), Community Health Centers (CHCs), and District Hospitals (DHs). The survey instrument focused on the availability of diagnostic tests as per the ICMR National Essential Diagnostics List (NEDL 2019) for NTDs (Table 1) (23). The study methodology was reviewed and approved by the Institutional Ethics Committee (Reference No. NEDL/2024–2025/IEC/FAED-01).

**Table 1.** ICMR National Essential Diagnostics List (NEDL), 2019–recommended diagnostic tests for selected neglected tropical diseases (NTDs) across different levels of public health facilities in India.

| S. NO. | Disease | Sub Centre |  | Primary Health Centre |  | Community Health Centre |  | District Hospital |  |
| --- | --- | --- | --- | --- | --- | --- | --- | --- | --- |
|  |  | Sample Type | Recommended Test | Sample Type | Recommended Test | Sample Type | Recommended Test | Sample Type | Recommended Test |
| 1 | Malaria | Whole Blood /Capillary Blood | Antigen based bivalent RDT for malaria | 1. Capillary blood<br>2. Whole Blood | 1.Peripheral smear for malaria parasite detection by Microscopy<br>2. Antigen based bivalent RDT for malaria | 1.Capillary blood<br>2. Whole Blood | 1.Peripheral smear for malaria parasite detection by Microscopy<br>2.Antigen based bivalent RDT for malaria | 1.Capillary blood<br>2.Whole Blood | 1.Peripheral smear for malaria parasite detection by Microscopy<br>2. Antigen based bivalent RDT for malaria |
| 2 | Filariasis | Capillary Blood (Peripheral blood thick smear) | Sample (prepared slide) to be collected and sent to PHC for microscopy | Capillary Blood (Peripheral blood thick smear) | Peripheral blood smear for filarial parasite detection (Thick smear) by Microscopy | Capillary Blood | Peripheral smear for parasite detection (Thick smear) by Microscopy | Capillary Blood | Peripheral smear for parasite detection (Thick smear) by Microscopy |
| 3 | Dengue | Not Recommended |  | Blood Serum for NS1 antigen and IgM antibody-based test. | Sample to be collected and sent to DH for ELISA | Blood Serum for NS1 antigen and IgM antibody-based test. | Sample to be collected and sent to DH for ELISA | Blood Serum | NS1 antigen and IgM antibody based test by ELISA |
| 4 | Chikungunya | Not Recommended |  | Not Recommended |  | Not Recommended |  | Blood Serum | IgM, Antibody test by ELISA |
| 5 | Japanese Encephelitis | Not Recommended |  | Blood Serum for IgM antibody based test. | Sample to be collected and sent to DH for ELISA | Blood Serum for IgM antibody based test. | Sample to be collected and sent to DH for ELISA | Blood Serum/CSF | IgM, Antibody test by ELISA |
| 6 | Leishmaniasis | Not Recommended |  | Blood Serum | rK39 test for Kala-Azar by RDT | Blood Serum | rK39 test for Kala-Azar by RDT | Blood Serum | rK39 test for Kala-Azar by RDT |
| 7 | HIV | Serum/ Plasma/ Whole blood | 1. Pre- test counselling for HIV screening.<br>2. HIV test (Antibodies 1/2) by RDT | Serum/ Plasma/ Whole blood | 1. Pre- test counselling for HIV screening.<br>2. HIV test (Antibodies 1/2) by RDT<br>3. ELISA (sample to be sent to DH and above) | Serum/ Plasma/ Whole blood | 1. Pre- test counselling for HIV screening.<br>2. HIV test (Antibodies 1/2) by RDT<br>3. ELISA (sample to be sent to DH and above) | Serum/ Plasma/ Whole blood | 1. Pre- test counselling for HIV screening.<br>2. HIV test (Antibodies 1/2) by RDT & ELISA<br>3. HIV screening in blood bank samples<br>4. CD4 count by Flow Cytometry |
| 8 | Helminthic Infections | Not Recommended |  | Stool | Stool routine examination including ova and parasite detection by Microscopy | Stool | Stool routine examination including ova and parasite detection by Microscopy | Stool | Stool routine examination including ova and parasite detection by Microscopy |

This table summarizes diagnostic tests recommended under the ICMR National Essential Diagnostics List (NEDL), 2019 for selected neglected tropical diseases, stratified by level of public health facility, including Sub-Centres (SCs), Primary Health Centres (PHCs), Community Health Centres (CHCs), and District Hospitals (DHs). Recommendations reflect the minimum essential diagnostic capacity expected at each facility level under the public health system.

#### Data collection

- ▪ Trained survey teams visited each facility between November 2023 and April 2025. They completed a standard diagnostic checklist (per NEDL 2019) to verify whether specific diagnostic tests were available on the day of the visit.
- ▪ Data were recorded on facility type, state, and test functionality.

#### National Neglected Tropical Diseases burden estimation

We collected national burden data for selected NTDs for the period 2021-2025 from publicly available sources, including NCVBDC, WHO, and other published reports.

### Data analysis

● We present percentages of facilities that perform specific diagnostic tests, stratified by state and facility level (DH vs. lower levels).
● We compare availability with ICMR NEDL recommendations.
● We present burden estimates (cases, prevalence, risk) for selected NTDs at the national level and, where data are available, by state.
● We examine whether states with higher burden have better diagnostic availability and identify gaps.

### Statistical Analysis

All statistical analyses were performed using **IBM SPSS Statistics version 26.0** (IBM Corp., Armonk, NY, USA). Descriptive statistics were calculated to summarise facility characteristics and diagnostic availability, including frequencies, proportions, means, and DRI. Continuous variables such as the Diagnostic Readiness Index (DRI) were compared across facility tiers using the **Kruskal–Wallis test**, followed by One Way-ANOVA.

### Calculation of Diagnostic Readiness Index (DRI)

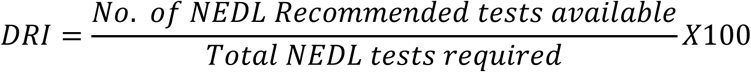

## Results

A total of 332 health-care facilities across seven states and one union territory were surveyed, comprising sub-centers (SCs; n=76), primary health centers (PHCs; n=152), community health centers (CHCs; n=80), and district hospitals (DHs; n=24). The study covered districts in Assam (Cachar, Goalpara, Hailakandi, Kamrup Metro, Sribhumi), Delhi (West Delhi, New Delhi), Haryana (Faridabad), Himachal Pradesh (Sirmaur), Manipur (Bishnupur, East Imphal, West Imphal), Rajasthan (Jaipur), Tripura (North Tripura, South Tripura, Unakoti, West Tripura), and Uttar Pradesh (Aligarh, Budaun, Bulandshahr, Gautam Budh Nagar, Kasganj). Facilities were assessed for the availability of diagnostic services for major neglected tropical diseases (NTDs) in accordance with the ICMR National Essential Diagnostics List (NEDL 2019). Overall, diagnostic readiness improved with increasing levels of care, although marked variations were observed across states and disease categories (Fig 1).

**Fig 1.**
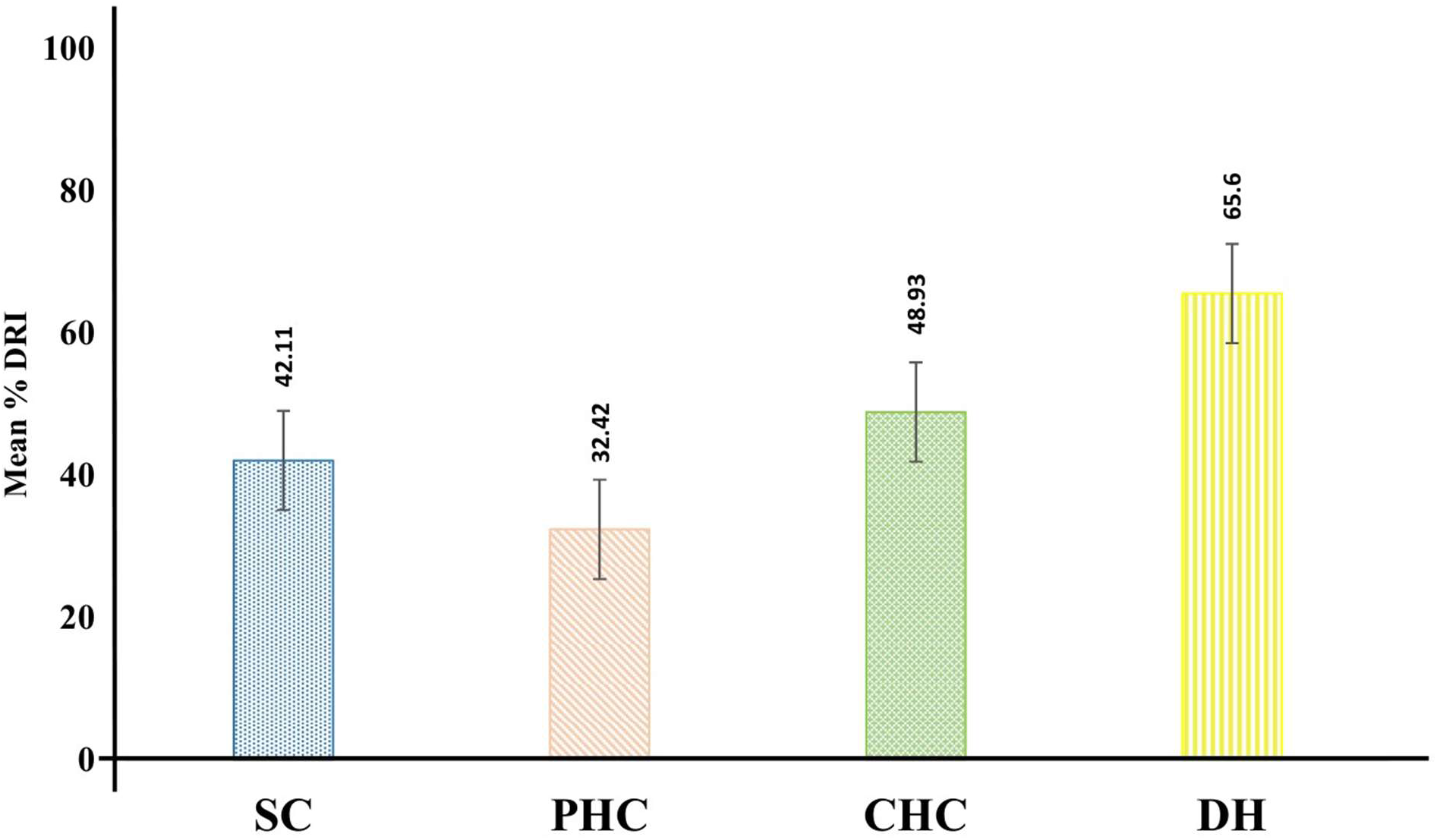
Facility-level comparison of mean DRI for selected NTDs across all surveyed state, (Each bar represents mean % DRI for all selected neglected tropical diseases (NTDs) across the surveyed health care facilities (Sub-Centers, PHCs, CHCs, and District Hospitals) aggregated across all surveyed states.

### Diagnostic Availability at Sub Centers (SCs)

At the Sub-Centre level, the mean percentage Diagnostic Readiness Index (DRI) for all NTDs and HIV was 42.11% (Fig 1), highest for malaria testing (81.58%), followed by HIV screening (39.47%) (Fig 2). In contrast, filariasis diagnostics showed extremely low readiness, with a DRI of only 5.26%, highlighting a major service gap at the peripheral tier (Fig 2). Among the surveyed regions, Tripura recorded the highest overall diagnostic readiness at SCs. Statistically, however, no significant difference in diagnostic availability was observed across states at the Sub-Centre level (Table 2 & Fig 3).

**Fig. 2.**
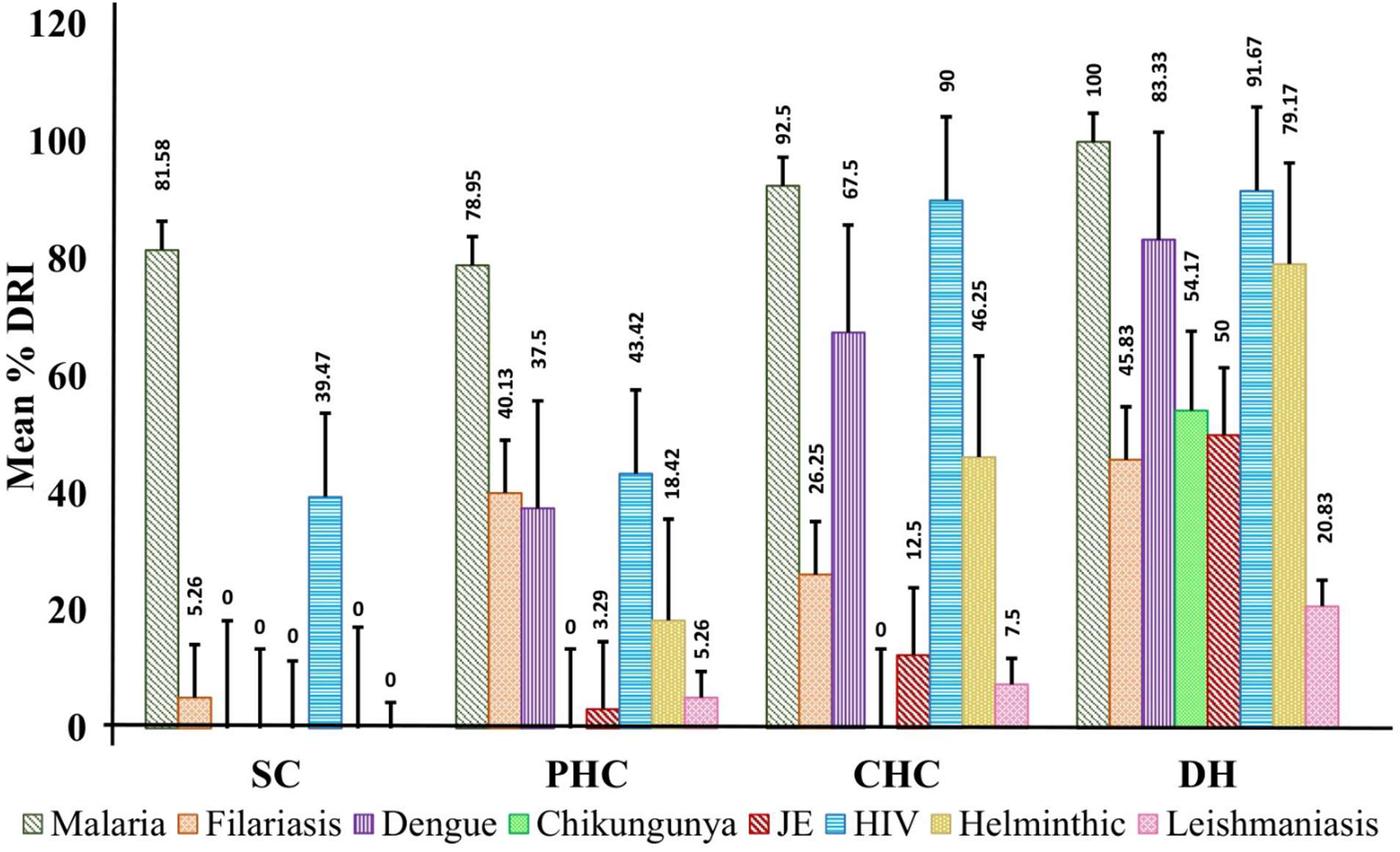
Facility-wise comparison of mean % DRI for each NTD and HIV across health care facilities in all surveyed states, (The figure presents a facility-wise comparison of mean % DRI for malaria, Filariasis, Dengue, Chikungunya, Japanese encephalitis, Helminthic infections, Leishmaniasis, and HIV. Mean DRI values are aggregated across all surveyed states for each level of health care facility (Sub-Centres, PHCs, CHCs, and District Hospitals). A value of **0%** indicates that either the survey was not conducted for that facility or that the respective diagnostic test was not available at the time of assessment).

**Fig 3.**
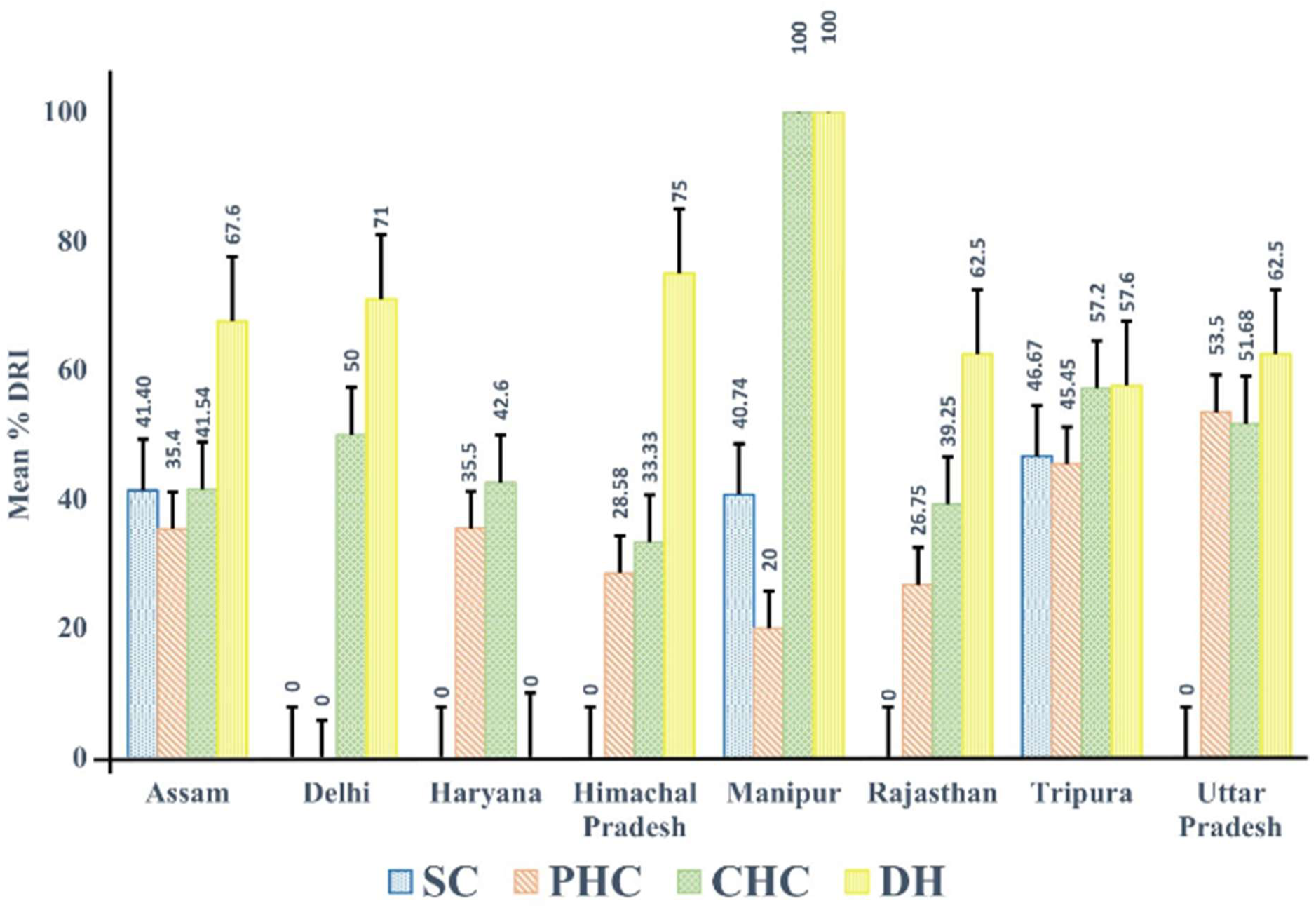
State-wise distribution of mean DRI for all NTDs and HIV across different health care facilities, {Each bar represents the mean percentage DRI for all neglected tropical diseases (NTDs) and HIV, aggregated across all surveyed health care facilities (Sub-Centres, PHCs, CHCs, and District Hospitals) within each surveyed state. A value of **0%** indicates that either the survey was not conducted for that facility or that the required diagnostic test was not available at the time of assessment}.

**Table. 2.** Diagnostic Readiness Index (DRI, %) for Neglected Tropical Diseases and HIV across public health-care facility levels (Sub-Centre [SC], Primary Health Centre [PHC], Community Health Centre [CHC], and District Hospital [DH]) in surveyed states of India.

| State & UT's |  | Assam | Delhi | Haryana | Himachal Pradesh | Manipur | Rajasthan | Tripura | Uttar Pradesh |
| --- | --- | --- | --- | --- | --- | --- | --- | --- | --- |
| Dengue | DRI at SC | NR | NR | NR | NR | NR | NR | NR | NR |
|  | DRI at PHC | 57.97 | NS | 50 | 0 | 20 | 0 | 81.82 | 100 |
|  | DRI at CHC | 50 | 50 | 66.67 | 0 | 100 | 0 | 100 | 91.18 |
|  | DRI at DH | 100 | 66.67 | NS | 100 | 100 | 100 | 100 | 57.14 |
| Chikungunya | DRI at SC | NR | NR | NR | NR | NR | NR | NR | NR |
|  | DRI at PHC | NR | NR | NR | NR | NR | NR | NR | NR |
|  | DRI at CHC | NR | NR | NR | NR | NR | NR | NR | NR |
|  | DRI at DH | 40 | 100 | NS | 100 | 100 | 100 | 0 | 57.14 |
| Japanese Encephalitis | DRI at SC | NR | NR | NR | NR | NR | NR | NR | NR |
|  | DRI at PHC | 2.9 | NS | 0 | 0 | 20 | 0 | 18.18 | 0 |
|  | DRI at CHC | 4.55 | 0 | 0 | 0 | 100 | 0 | 20 | 11.76 |
|  | DRI at DH | 80 | 66.67 | NS | 100 | 100 | 0 | 40 | 28.57 |
| Malaria | DRI at SC | 80.65 | NS | NS | NS | 66.66 | NS | 100 | NS |
|  | DRI at PHC | 63.77 | NS | 100 | 100 | 20 | 100 | 81.82 | 100 |
|  | DRI at CHC | 77.27 | 100 | 100 | 83.33 | 100 | 100 | 100 | 100 |
|  | DRI at DH | 100 | 100 | 100 | 100 | 100 | 100 | 100 | 100 |
| HIV | DRI at SC | 40.32 | NS | NS | NS | 33.33 | NS | 40 | NS |
|  | DRI at PHC | 68.12 | NS | 100 | 0 | 80 | 87.5 | 90.91 | 100 |
|  | DRI at CHC | 86.36 | 100 | 100 | 33.33 | 100 | 100 | 100 | 97.06 |
|  | DRI at DH | 60 | 100 | NS | 100 | 100 | 100 | 100 | 100 |
| Leishmaniasis | DRI at SC | NR | NR | NR | NR | NR | NR | NR | NR |
|  | DRI at PHC | 7.25 | NS | 0 | 0 | 0 | 0 | 18.18 | 25 |
|  | DRI at CHC | 4.55 | 0 | 0 | 0 | 100 | 0 | 20 | 0 |
|  | DRI at DH | 20 | 0 | NS | 0 | 100 | 0 | 0 | 42.86 |
| Helminthic Infections | DRI at SC | NR | NR | NR | NR | NR | NR | NR | NR |
|  | DRI at PHC | 36.23 | NS | 0 | 0 | 0 | 0 | 18.18 | 0 |
|  | DRI at CHC | 31.82 | 100 | 33.33 | 0 | 100 | 75 | 20 | 55.88 |
|  | DRI at DH | 80 | 100 | 100 | 100 | 100 | 100 | 20 | 100 |
| Filariasis | DRI at SC | 3.23 | NS | NS | NS | 22.22 | NS | 0 | NS |
|  | DRI at PHC | 11.59 | NS | 0 | 100 | 0 | 0 | 9.09 | 50 |
|  | DRI at CHC | 36.36 | 0 | 0 | 83.33 | 100 | 0 | 40 | 5.88 |
|  | DRI at DH | 60 | 33.33 | NS | 0 | 100 | 0 | 100 | 14.29 |
|  | *p* Value | 0.035 | 0.068 | 0.057 | 0.548 | 0.867 | 0.042 | 0.011 | 0.03 |
| <sup>a</sup> NR: Not Recommended as per ICMR NEDL 2019<br><sup>b</sup> NS: Survey not done |  |  |  |  |  |  |  |  |  |

### Diagnostic Availability at Primary Health Centers (PHCs)

The mean percentage DRI for PHC across all selected NTDs and HIV was 32.42% (Fig 1). At the PHC level, the Diagnostic Readiness Index (DRI) indicated that malaria had the highest availability (78.95%), followed by HIV (43.42%) and filariasis (40.13%). DRI was much lower lower for dengue (37.50%), helminthic infections (18.42%), and abysmal for leishmaniasis (5.26%), and Japanese encephalitis (JE) (3.29%) (Fig 2).

State-wise assessment (Table 2) revealed that Himachal Pradesh, Rajasthan, and Uttar Pradesh demonstrated full readiness (DRI: 100%) for malaria diagnosis, followed by Tripura (81.82%), Haryana (80%), and Assam (63.77%), while Manipur showed the lowest readiness (20%). For dengue, Uttar Pradesh achieved the highest DRI (100%), followed by Tripura (81.82%) and Assam (57.97%), whereas Himachal Pradesh and Rajasthan reported no readiness for dengue diagnostics.

HIV diagnostic readiness was highest in Uttar Pradesh (100%), followed by Tripura (90.91%), Rajasthan (87.5%), Haryana (80%), Manipur (80%), and Assam (53.62%) but absent in Himachal Pradesh. JE diagnostic readiness remained limited, with maximum availability in Manipur (20%) and Tripura (18.18%). Filariasis readiness was highest in Himachal Pradesh (100%) and Uttar Pradesh (50%), but absent in Haryana, Manipur, and Rajasthan. The diagnosis of helminthic infection showed the highest readiness in Assam (36.23%) and Tripura (18.18%), with no services available in several states. Leishmaniasis diagnostics were most available in Uttar Pradesh (25%), followed by Tripura (18.18%) and Assam (7.25%).

### Diagnostic Availability at Community Health Centers (CHCs)

The mean percentage DRI of CHC for all NTDs and HIV was 48.93% (Fig 1), Across CHCs, the Diagnostic Readiness Index showed substantial variation across diseases and states. Malaria demonstrated the highest readiness, with an overall availability of 92.50%, and full readiness (100%) in Delhi, Haryana, Manipur, Rajasthan, Tripura, and Uttar Pradesh; slightly lower levels were noted in Assam (77.27%) and Himachal Pradesh (83.33%). HIV diagnostic readiness was similarly high at 90%, with most states achieving 100% except Himachal Pradesh (33.33%), Assam (86.36%), and Uttar Pradesh (97.06%).

For dengue, the diagnostic readiness index was 67.50%, reaching full availability in Manipur and Tripura, and remaining high in Uttar Pradesh (91.18%), moderate in Haryana (66.67%), and low in Assam and Delhi (50%), with no readiness in Himachal Pradesh and Rajasthan. Diagnostic testing for helminthic infections showed moderate readiness (46.25%), with complete availability in Delhi and Manipur, followed by Rajasthan (75%), Uttar Pradesh (55.88%), Haryana (33.33%), Assam (31.82%), and Tripura (20%). Filariasis diagnostic readiness was low at 26.25%, though high in Manipur (100%) and Himachal Pradesh (83.33%); moderate in Tripura (40%) and Assam (36.36%); minimal in Uttar Pradesh (5.88%), and absent in Delhi, Haryana, and Rajasthan (0%). Japanese encephalitis (JE) exhibited limited readiness (12.50%), with full availability only in Manipur, and very low levels in Tripura (20%), Uttar Pradesh (11.76%), and Assam (4.55%); no readiness was observed in Delhi, Haryana, Himachal Pradesh, and Rajasthan. Diagnostic tests for leishmaniasis showed the lowest readiness (7.50%), with availability only in Manipur (100%), Tripura (20%), and Assam (4.55%), and absent in most other states (Table 2 & Fig 2).

### Diagnostic Availability at District Hospitals (DHs)

At the District Hospital (DH) level, the mean Diagnostic Readiness Index (DRI) was highest (67.19%) among all healthcare tiers (Fig 1). Malaria diagnosis achieved a perfect DRI (100%) across all surveyed DHs. HIV testing showed a strong DRI of 91.67%, with complete readiness in Delhi, Himachal Pradesh, Manipur, Rajasthan, Tripura, and Uttar Pradesh. Dengue diagnostics achieved an overall DRI of 83.33%, with full readiness in most states except Delhi (66.67%) and Uttar Pradesh (57.14%). Helminthic infection diagnostics showed a DRI of 79.17%, with complete readiness in Delhi, Himachal Pradesh, Manipur, Rajasthan, and Uttar Pradesh; availability was moderate in Assam (80%) and limited in Tripura (20%). Chikungunya testing—recommended exclusively at the DH level—exhibited a DRI of 54.17%, with full readiness in Delhi, Himachal Pradesh, Manipur, and Rajasthan; moderate readiness in Uttar Pradesh (57.14%) and Assam (40%); and no availability in Tripura. Japanese encephalitis (JE) diagnostics had a DRI of 50%, with Himachal Pradesh and Manipur achieving full readiness, followed by Assam (80%), Delhi (66.67%), Tripura (40%), and Uttar Pradesh (28.57%). Filariasis testing showed a DRI of 45.83%, with the highest readiness in Tripura and Manipur (100%). Diagnostic services for leishmaniasis had the lowest DRI (20.83%), although Manipur exhibited full readiness, followed by Uttar Pradesh (42.86%) and Assam (20%), with no availability in the remaining surveyed states (Table 2 & Fig 2).

### Diagnostic Readiness Index at the District Level

District-level analysis of mean diagnostic readiness index (DRI), aggregating all public health facilities (SCs, PHCs, CHCs & DHs), revealed substantial inter-district and disease-specific heterogeneity (Table 3 & Fig 4). Malaria demonstrated the highest overall readiness (mean DRI: 84.34%), with >90% DRI in most districts, although markedly lower readiness was observed in Cachar, East Imphal, and North Tripura. Chikungunya (61.90%) and HIV (57.23%) showed moderate overall readiness, with several districts achieving 100% DRI. Dengue readiness was variable (mean DRI: 40.36%), ranging from complete readiness in selected districts to ≤80% in many others. In contrast, diagnostic readiness for filariasis (29.22%), helminthic infections (25.30%), Japanese encephalitis (8.13%), and leishmaniasis (5.72%) was uniformly poor across districts, despite isolated high-performing sites. These findings highlight critical gaps in district-level diagnostic preparedness for neglected tropical diseases across different health care facilities, particularly beyond malaria and select arboviral infections.

**Fig 4.**
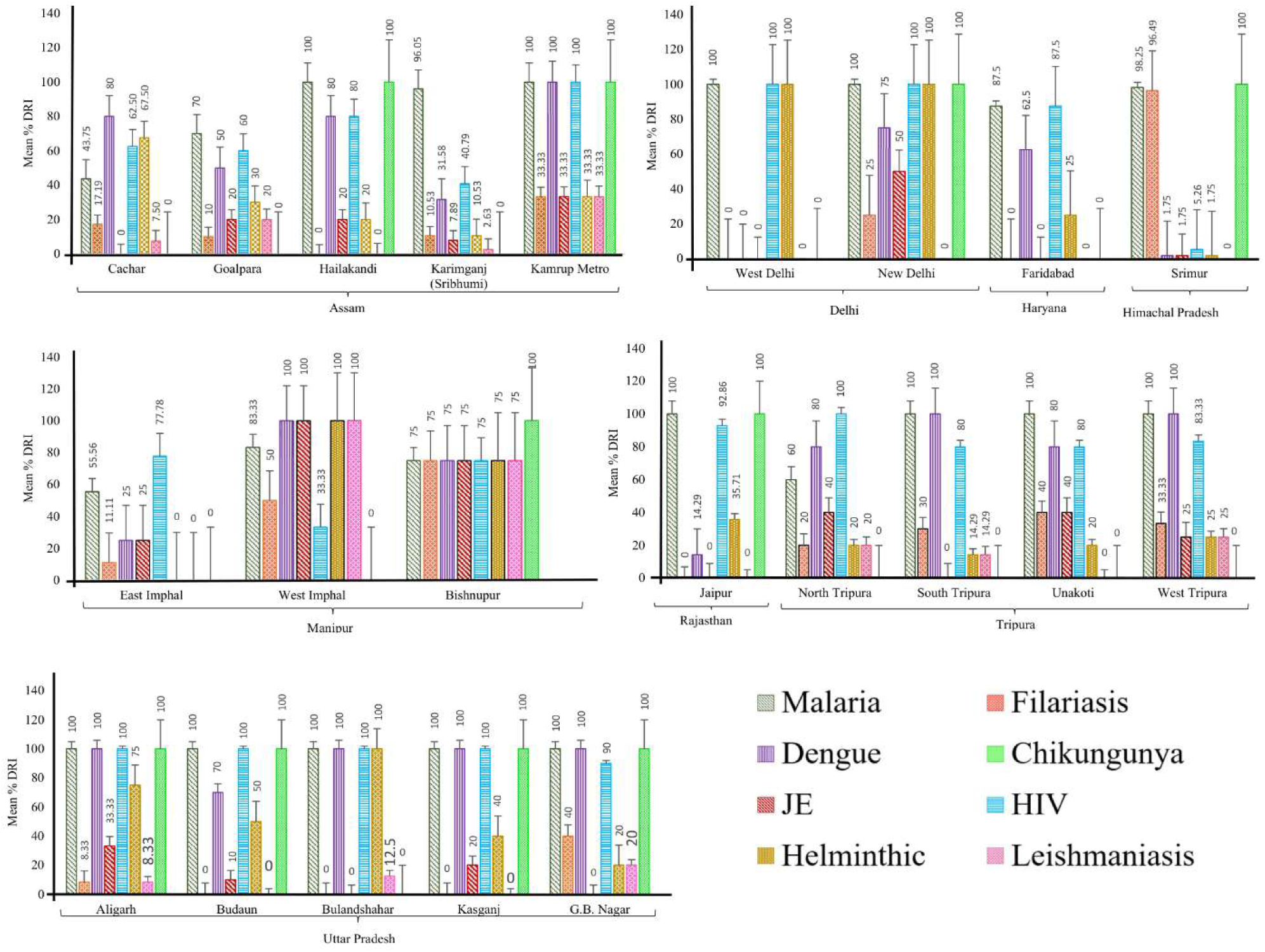
Mean DRI of District health care facilities (SC, PHC, CHC & DH) across different states of India (a) Mean DRI of health care facilities across surveyed districts of Assam. (b) Mean DRI of health care facilities across surveyed districts of Delhi, Haryana & Himachal Pradesh. (c) Mean DRI of health care facilities across surveyed districts of Manipur. (d) Mean DRI of health care facilities across surveyed districts of Rajasthan & Tripura. (e) Mean DRI of health care facilities across surveyed districts of Uttar Pradesh.

**Table 3.**
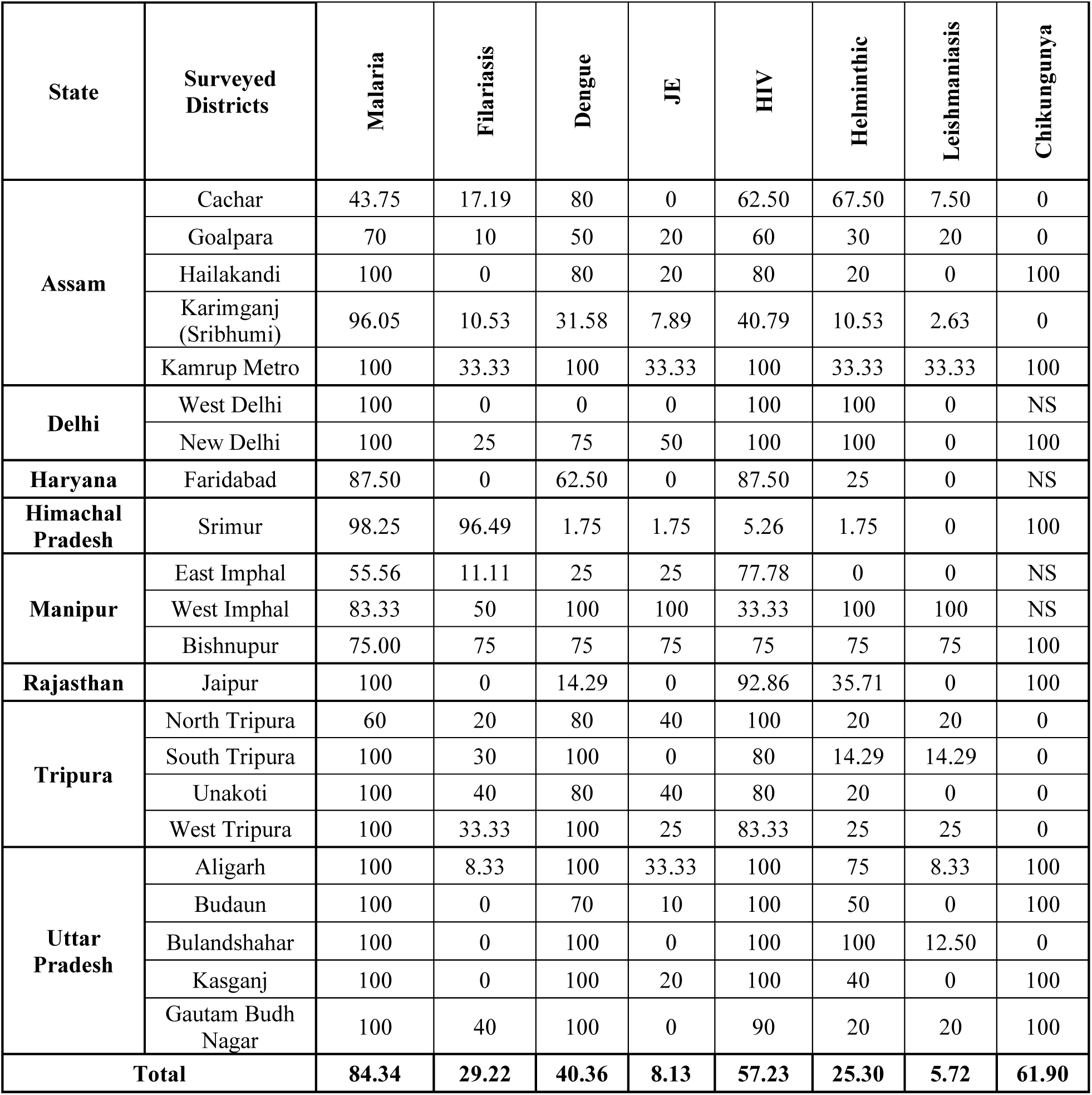
Aggregated percentage Diagnostic Readiness Index (DRI, %) of district-level public health-care facilities—Sub-Centres (SCs), Primary Health Centres (PHCs), Community Health Centres (CHCs), and District Hospitals (DHs)—for neglected tropical diseases (NTDs) and HIV across surveyed districts in participating states.

### State-wise comparison of diagnostic availability

Overall diagnostic availability varied markedly across the surveyed states (Fig 5). Delhi showed higher overall diagnostic availability, with multiple essential diagnostics consistently available across disease categories, indicating stronger diagnostic coverage than in most other states. Manipur also showed relatively high overall availability, reflecting broader access to diagnostics for both vector-borne and other neglected tropical diseases. Uttar Pradesh and Rajasthan exhibited moderate overall diagnostic availability, with good coverage of malaria, dengue, chikungunya, and HIV, but notable gaps in coverage for filariasis, Japanese encephalitis, helminthic infections, and leishmaniasis. Assam showed lower overall diagnostic availability compared with Delhi and Manipur, with availability concentrated primarily for malaria, dengue, chikungunya, and HIV, and limited coverage for other NTDs. Tripura demonstrated mixed availability, with adequate access to malaria, dengue, filariasis, and HIV diagnostics, but the absence of diagnostics for several other NTDs, resulting in lower overall availability. Haryana showed comparatively limited overall diagnostic availability, largely restricted to malaria, dengue, and HIV. Himachal Pradesh demonstrated uneven overall availability, with good access to diagnostics for malaria, filariasis, and chikungunya but limited or absent availability for several other priority NTDs.

**Fig 5.**
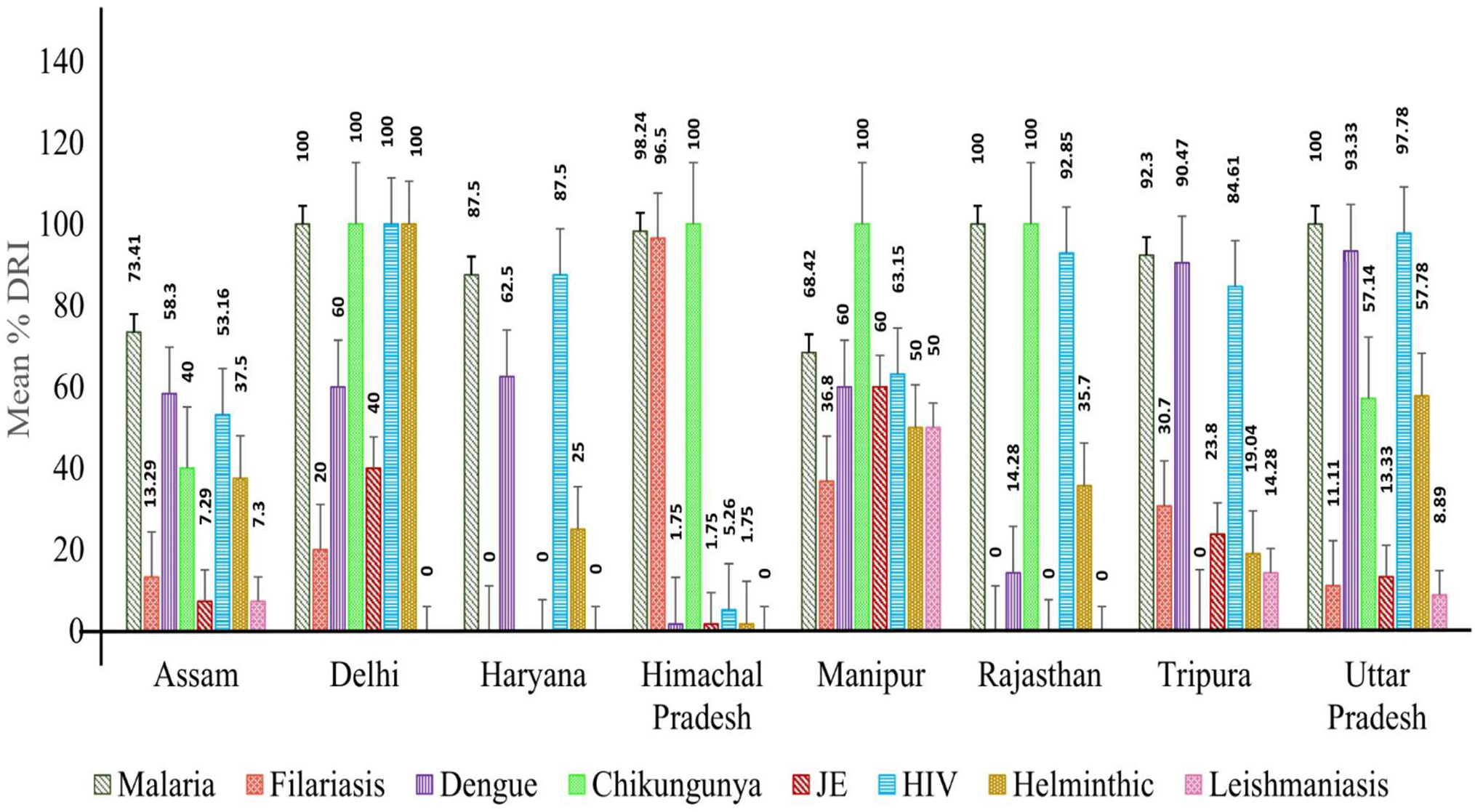
State-wise comparison of mean percentage DRI for individual NTDs and HIV across different states. (The figure depicts the state-wise mean percentage DRI for each selected neglected tropical disease—Malaria, Filariasis, dengue, chikungunya, Japanese encephalitis, Helminthic infections, and Leishmaniasis—as well as HIV, calculated by aggregating data from all surveyed health care facilities (Sub-Centres, PHCs, CHCs, and District Hospitals) within each state. A value of **0%** indicates that either the survey was not conducted for the respective facility or that the specific diagnostic test was not available at the time of assessment).

### Burden of Neglected Tropical Diseases and HIV in India

National surveillance data indicate a substantial burden of neglected tropical diseases and HIV in India during the most recent reporting period. In 2024, malaria accounted for 255,500 confirmed cases and 86 deaths, remaining one of the most widely reported vector-borne diseases. Dengue represented a major epidemic-prone illness, with 233,529 cases and 297 deaths, while Japanese encephalitis (JE) contributed 1,472 cases and 105 deaths, reflecting continued focal transmission and high fatality. Chikungunya was reported to have caused 17,930 cases nationally in 2024, indicating sustained arboviral circulation. Visceral leishmaniasis (kala-azar) persisted at lower but ongoing levels, with 449 cases and 9 deaths reported in 2024 (Fig 6). In addition, lymphatic filariasis continues to contribute a substantial chronic disease burden, with 621,178 lymphoedema cases and 127,100 hydrocele cases reported in 2023 (46). Beyond NTDs, HIV remains a major public health condition, with an estimated 2.5 million people living with HIV (PLHIV) in 2023, alongside 66,450 new infections and 35,870 AIDS-related deaths (47).

**Fig. 6.**
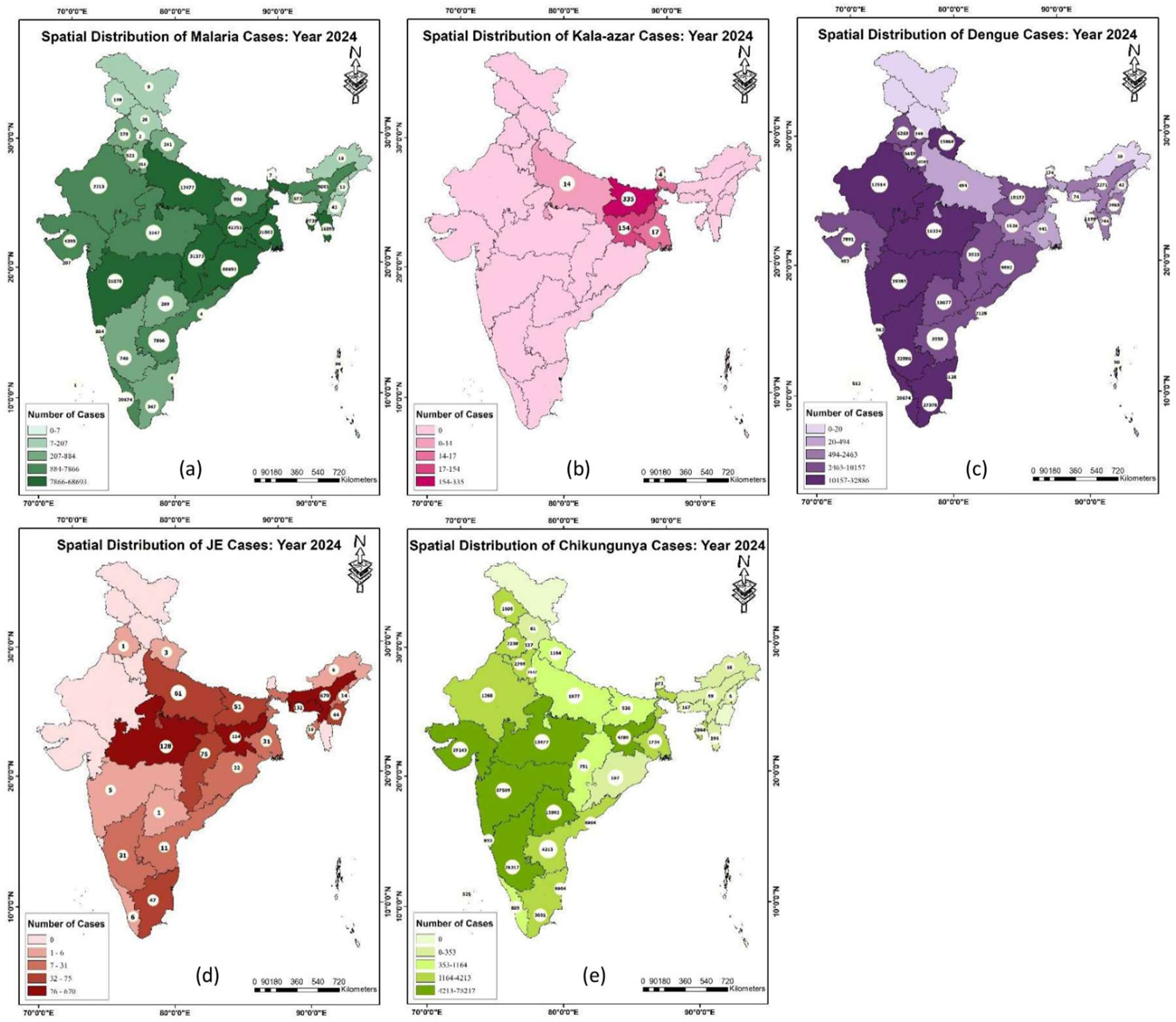
Spatial distribution of Vector-Borne Diseases across different states of India-2024 (Maps were generated using ArcGIS pro software; Data source: NCVBDC). (a)Spatial distribution of Malaria cases across different Indian states. (b) Spatial distribution of Visceral Leishmaniasis (Kala Azar) cases across different Indian states. (c) Spatial distribution of Dengue cases across different Indian states. (d) Spatial distribution of JE cases across different Indian states. (e) Spatial distribution Chikungunya cases across different Indian states.

## Discussion

This multi-state facility-based assessment demonstrates a pronounced tiered gradient in diagnostic readiness across India’s public health system, with the highest availability consistently observed at district hospitals, intermediate readiness at Community Health Centers (CHCs), and substantial deficiencies at Primary Health Centers (PHCs) and Sub-Centers. This hierarchical pattern reflects long-standing disparities in infrastructure, laboratory capacity, workforce skills, procurement systems, and programme prioritization across health-system tiers (24). While diagnostic readiness shows strong concordance with disease burden and programmatic emphasis for malaria and HIV, this alignment is weak, inconsistent, or absent for most neglected tropical diseases (NTDs), including Dengue, Chikungunya, Lymphatic Filariasis (LF), Japanese encephalitis (JE), visceral Leishmaniasis (VL), and Helminthic infections. These findings underscore the critical role of sustained policy commitment and financing in shaping diagnostic capacity, while also revealing persistent operational and implementation gaps that undermine early detection and effective surveillance for several NTDs in India.

Malaria exhibited consistently high diagnostic readiness across all health-system levels, including SCs and PHCs. This reflects decades of sustained national prioritization, supported by validated rapid diagnostic tests (RDTs), standardized procurement mechanisms, quality assurance systems, and continuous workforce training. The integration of antigen-based RDTs with microscopy in routine service delivery has enabled the widespread decentralization of malaria diagnosis, facilitating early case detection and prompt treatment initiation even in remote settings. National Programme reports further confirm the near-universal availability of malaria diagnostics at peripheral facilities, reinforcing malaria’s unique position among vector-borne diseases in India. Facility-based assessments from multiple regions similarly report high diagnostic availability for malaria at SCs and PHCs, in stark contrast to limited diagnostic capacity for other NTDs (25–27). The state-wise patterns observed in the present study—showing high or complete readiness in Himachal Pradesh, Rajasthan, and Uttar Pradesh, moderately lower readiness in Tripura, Haryana, and Assam, and comparatively weaker coverage in Manipur—highlight how sustained programmatic investment can achieve widespread diagnostic availability while also exposing regional inequities. Overall, the malaria Programme illustrates how strong policy commitment, dedicated financing, and systematic implementation can successfully embed decentralized diagnostics across health-system tiers. Replicating these elements for other NTDs could substantially improve diagnostic readiness and surveillance capacity nationwide.

Diagnostic readiness for dengue and other arboviral diseases in this study was uneven, with most testing occurring at higher-level health facilities. Although national guidelines recommend that sample collection for Dengue and Japanese encephalitis be initiated at PHCs and CHCs with referral to designated laboratories for confirmatory testing, the low readiness at the primary care level suggests persistent implementation bottlenecks that may weaken surveillance, delay diagnosis, and compromise outbreak response. Current dengue surveillance in India is embedded within the National Vector Borne Disease Control Programme’s sentinel surveillance network, which consists of hundreds of sentinel hospitals and a smaller number of referral laboratories where confirmatory IgM ELISA testing is conducted; the distribution of these sentinel sites varies substantially between states and remains limited compared to the overall burden of disease (28). Point-of-care NS1/IgM rapid diagnostic tests have been evaluated in Indian clinical settings and have shown reasonable diagnostic performance, with one study reporting sensitivities of approximately 89.4% and specificities of 93.8% for combined NS1 and IgM detection across all four dengue serotypes, compared with RT-PCR and ELISA reference methods (29). However, other comparative assessments indicate that NS1 and IgM rapid diagnostic tests differ in sensitivity and specificity from ELISA, and their reliability as standalone diagnostics is often lower, particularly at later stages of illness or in peripheral health settings (30). These findings underscore the requirement for confirmatory ELISA or molecular testing at well-equipped laboratories, particularly for surveillance quality and accurate case classification in endemic regions.

Despite the potential utility of rapid tests, multiple studies and surveillance reviews indicate limited integration of dengue diagnostic tools into routine primary care services in India, with most advanced diagnostics occurring at CHCs, district hospitals, and referral laboratories rather than at the PHC level (28). Consistent with this evidence base, the present study found substantially lower dengue diagnostic readiness at PHCs, moderate improvement at CHCs, and the highest availability at district hospitals. State-level variation was evident, with comparatively higher preparedness at PHCs in Uttar Pradesh, moderate readiness in Tripura and Assam, and absence of dengue diagnostics at the PHC level in Himachal Pradesh and Rajasthan. Although readiness increased at CHCs and district hospitals, inter-state gaps persisted, reinforcing the pattern of tiered diagnostic capacity concentrated at secondary and tertiary facilities documented in surveillance system analyses.

Chikungunya diagnostic readiness, recommended primarily at district hospital levels, showed considerable inter-state variation. While some states demonstrated full preparedness, others showed moderate or no readiness, reflecting disparities in laboratory infrastructure, workforce capacity, and resource allocation. The limited decentralization of chikungunya diagnostics to peripheral facilities may delay case confirmation and impede timely outbreak response, particularly in endemic and outbreak-prone regions. These findings align with previous assessments showing that chikungunya surveillance in India relies heavily on district and tertiary laboratories, with suboptimal integration into peripheral health facilities (31,32). Strengthening district hospital capacity, streamlining referral pathways, and expanding laboratory networks—particularly during outbreaks—are essential to improving the timeliness of detection, diagnosis, and response.

Despite the proven effectiveness of rK39 antigen-based rapid diagnostic tests (RDTs) for Visceral Leishmaniasis (VL), diagnostic readiness at PHC and CHC levels remained markedly limited across most affected districts in the present study. PHC-level readiness was extremely low, with comparatively higher availability observed only in Uttar Pradesh, followed by Tripura and Assam, while CHC-level readiness was even more restricted; district hospitals demonstrated relatively better, though still suboptimal, coverage. These gaps are unlikely to be attributable to diagnostic limitations, as rK39 RDTs have consistently shown high sensitivity and specificity under field conditions and are recommended as the primary diagnostic tool for VL in the Indian subcontinent, including in low-incidence and elimination settings (33,34). Instead, the observed deficiencies point to operational constraints, including irregular procurement, inadequate training for frontline health workers, and weaknesses in supply chain management. Evaluations of India’s VL elimination Programme have similarly reported that rK39 testing remains disproportionately concentrated at higher-level facilities despite national guidelines advocating early diagnosis and treatment at peripheral health centers (35). Empirical evidence further indicates that delays in VL diagnosis contribute to ongoing transmission and poorer clinical outcomes, underscoring the importance of decentralized diagnostic access. Expanding rK39 availability at PHCs and CHCs, alongside strengthened training, supervision, and logistics support, is therefore critical to improving early case detection and accelerating VL elimination efforts (33–35).

The mismatch between lymphatic Filariasis (LF) burden and diagnostic readiness was particularly pronounced in the present study. Diagnostic availability at SCs was extremely low, indicating a critical gap at the most peripheral tier of the health system, whereas readiness at PHCs and CHCs varied substantially across states. District hospitals consistently demonstrated the highest availability, reflecting the concentration of microscopy capacity and trained personnel at higher levels of care (36). Similar patterns have been documented in national and subnational assessments, which show that LF diagnostics remain weakly integrated into routine peripheral health services, particularly in post–mass drug administration (MDA) settings where surveillance intensity often declines (37,38). Although national guidelines recommend night blood collection at subcenters, with referral for microscopic confirmation, persistent constraints in laboratory infrastructure, trained human resources, supervision, and referral mechanisms continue to limit effective implementation (36). Recent evaluations have further highlighted challenges in maintaining microscopy quality in peripheral laboratories due to declining caseloads, competing workloads, and limited quality assurance, underscoring the need to strengthen diagnostic capacity beyond district-level facilities (36–38).

JE diagnostic readiness was among the lowest across all diseases assessed, despite its high case fatality and risk of long-term neurological sequelae. Diagnostic capacity at PHCs—where decentralized sample collection is recommended—was extremely limited, with meaningful readiness observed in only a few states. CHC-level preparedness was similarly poor, while district hospitals showed variable but still insufficient coverage. These findings reflect broader challenges in India’s JE surveillance system, including weak sample-collection systems, inadequate training, and logistical barriers at the peripheral level. Field studies from northeast India further demonstrate overlapping transmission of JE, dengue, and chikungunya, yet routine diagnostic pathways at peripheral facilities often fail to capture this complexity (39,40). Strengthening decentralized sample collection, referral systems, and integrated fever surveillance is therefore essential.

HIV diagnostic readiness was relatively high at PHC, CHC, and district hospital levels, reflecting the sustained impact of India’s national HIV Programme, standardized testing algorithms, and widespread availability of rapid diagnostic tests integrated into routine primary care. Decentralized HIV testing has been shown to improve early diagnosis, timely treatment initiation, and reduction of onward transmission, supporting the readiness patterns observed in this study (41,42). In contrast, diagnostic availability at SCs remained limited, consistent with national guidelines that prioritise HIV testing at higher-tier facilities, but also indicative of ongoing gaps in referral linkages, community outreach, and early case identification. Marked interstate variation was evident, with several states achieving near-complete preparedness, whereas others, including Himachal Pradesh, lagged behind. Similar regional disparities in HIV diagnostic coverage have been documented in facility-based and programmatic assessments across India, particularly in hard-to-reach and low-prevalence settings where testing uptake remains suboptimal (43,44). Strengthening SCs referral mechanisms and community-based testing linkages could further enhance equitable access and continuity of HIV care.

Diagnostic readiness for helminthic infections was higher than for many NTDs but remained uneven across tiers. Peripheral facilities continued to face gaps, consistent with evidence that helminth diagnostics remain underprioritized in routine surveillance (45,46). WHO and national guidelines emphasize the need for decentralized, quality-assured diagnostics to support integrated NTD control (47,48).

## Conclusions

The findings highlight persistent inequities in the availability of essential diagnostics for neglected tropical diseases in India, with readiness only partially aligned with the disease burden. While malaria diagnostics illustrate the impact of sustained programmatic investment, diagnostic capacity for other high-burden NTDs—including dengue, filariasis, Japanese encephalitis, and leishmaniasis remains inadequate at Sub-Centers and Primary Health Centers. This misalignment reflects systemic gaps in the implementation of the ICMR National Essential Diagnostics List and contributes to delayed detection, underreporting, and ongoing transmission. Strengthening burden-responsive planning, decentralizing point-of-care diagnostics, and improving supply-chain and surveillance integration are critical to enhance early diagnosis, treatment, equity in access to care, and progress toward national NTD control and elimination goals.

## Data Availability

All the relevant information has been reported in the manuscript.

## Acknowledgments

The authors sincerely thank all the staff and facilities who generously agreed to participate in this study and supported the data collection process. We are especially grateful to our advisory board for their valuable guidance and constructive input throughout the study. We also extend our heartfelt appreciation to the Ethics Committee for their continuous guidance and oversight, which ensured the ethical and systematic conduct of data collection.

## Notes

### Competing Interest Statement

The authors have declared no competing interest.

### Funding Statement

The author(s) received no specific funding for this work.

### Author Declarations

The study methodology was reviewed and approved by the FAED's Institutional Ethics Committee (Reference No. NEDL/2024-2025/IEC/FAED-01).

